# Deep Learning for Contrast Enhanced Mammography - a Systematic Review

**DOI:** 10.1101/2024.05.13.24307271

**Authors:** Vera Sorin, Miri Sklair-Levy, Benjamin S. Glicksberg, Eli Konen, Girish N. Nadkarni, Eyal Klang

## Abstract

**Background/Aim:** Contrast-enhanced mammography (CEM) is a relatively novel imaging technique that enables both anatomical and functional breast imaging, with improved diagnostic performance compared to standard 2D mammography. The aim of this study is to systematically review the literature on deep learning (DL) applications for CEM, exploring how these models can further enhance CEM diagnostic potential.

**Methods:** This systematic review was reported according to the PRISMA guidelines. We searched for studies published up to April 2024. MEDLINE, Scopus and Google Scholar were used as search databases. Two reviewers independently implemented the search strategy.

**Results:** Sixteen relevant studies published between 2018 and 2024 were identified. All studies but one used convolutional neural network models. All studies evaluated DL algorithms for classification of lesions at CEM, while six studies also assessed lesion detection or segmentation. In three studies segmentation was performed manually, two studies evaluated both manual and automatic segmentation, and ten studies automatically segmented the lesions.

**Conclusion:** While still at an early research stage, DL can improve CEM diagnostic precision. However, there is a relatively small number of studies evaluating different DL algorithms, and most studies are retrospective. Further prospective testing to assess performance of applications at actual clinical setting is warranted.

**Graphic Abstract:** 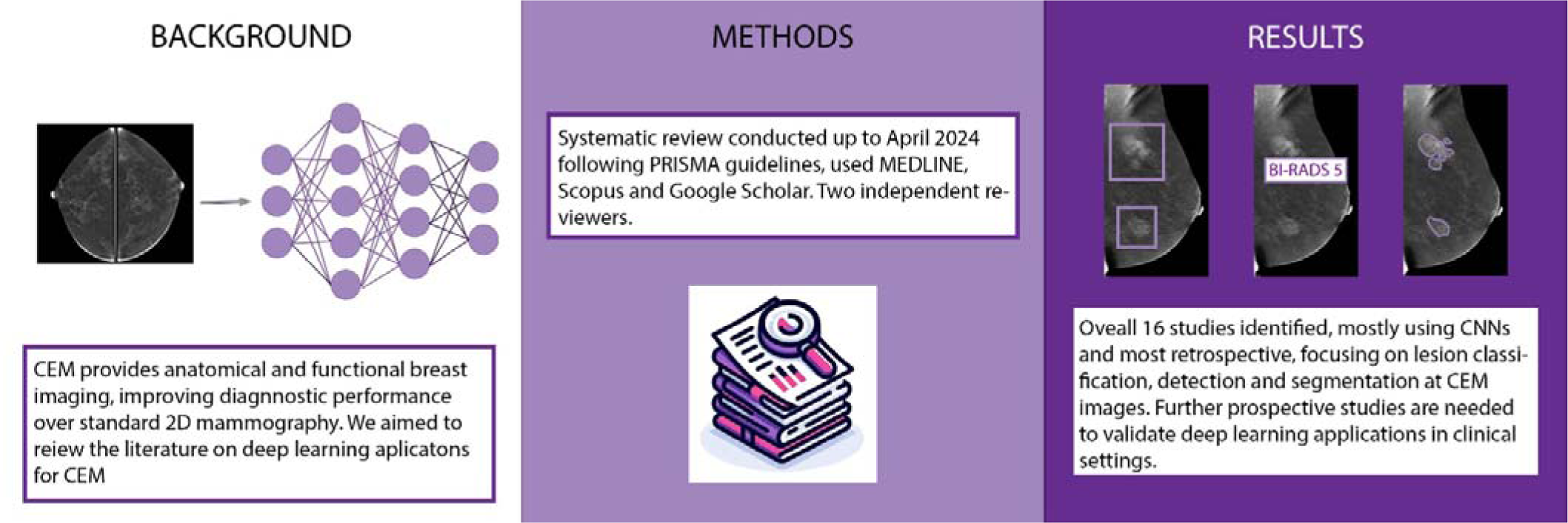

## Introduction

Despite being the cornerstone of breast imaging, 2D standard mammography faces well-known diagnostic challenges (1–3). Contrast-enhanced mammography (CEM) is an advanced imaging technique that enables both anatomical and functional breast imaging (4). The technique enhances the visibility of neo-vascularity in breast cancer. It illustrates pathologic enhancement in breast malignancies, sometimes even before the development of a distinct breast mass (5). Comparative studies have demonstrated CEM’s superior diagnostic performance over conventional mammography and a comparable sensitivity to breast MRI(6).

Recent advancements in deep learning (DL) have revolutionized computer vision in radiology (7). DL algorithms are increasingly applied to enhance the segmentation, detection, and classification of pathologies across various imaging modalities, including CEM (8). However, the integration of DL in CEM is still nascent, with significant potential to improve diagnostic accuracy, and enhance radiologists’ workflow.

Our study aims to systematically review the literature on DL applications for CEM, exploring how these models can further enhance its diagnostic potential.

## Methods

### Literature Search

We systematically searched for papers published up to April 9, 2024. MEDLINE, Scopus and Google Scholar were used as databases. The search query was: “((deep learning[Title/Abstract]) OR (CNN[Title/Abstract]) OR (AI[Title/Abstract]) OR (artificial intelligence[Title/Abstract]) OR (neural network[Title/Abstract])) AND ((CEM[Title/Abstract]) OR (CESM[Title/Abstract]) OR (contrast enhanced mammography[Title/Abstract]) OR (contrast mammography[Title/Abstract]) OR (DEM[Title/Abstract]))”.

### Eligibility Criteria

We included full publications indexed in PubMed, evaluating DL for automatic analysis of CEM images. We excluded non-English papers, non-original manuscripts, and papers that did not evaluate DL algorithms. **Figure 1** presents a flow diagram of the screening and inclusion process.

**Figure 1.**
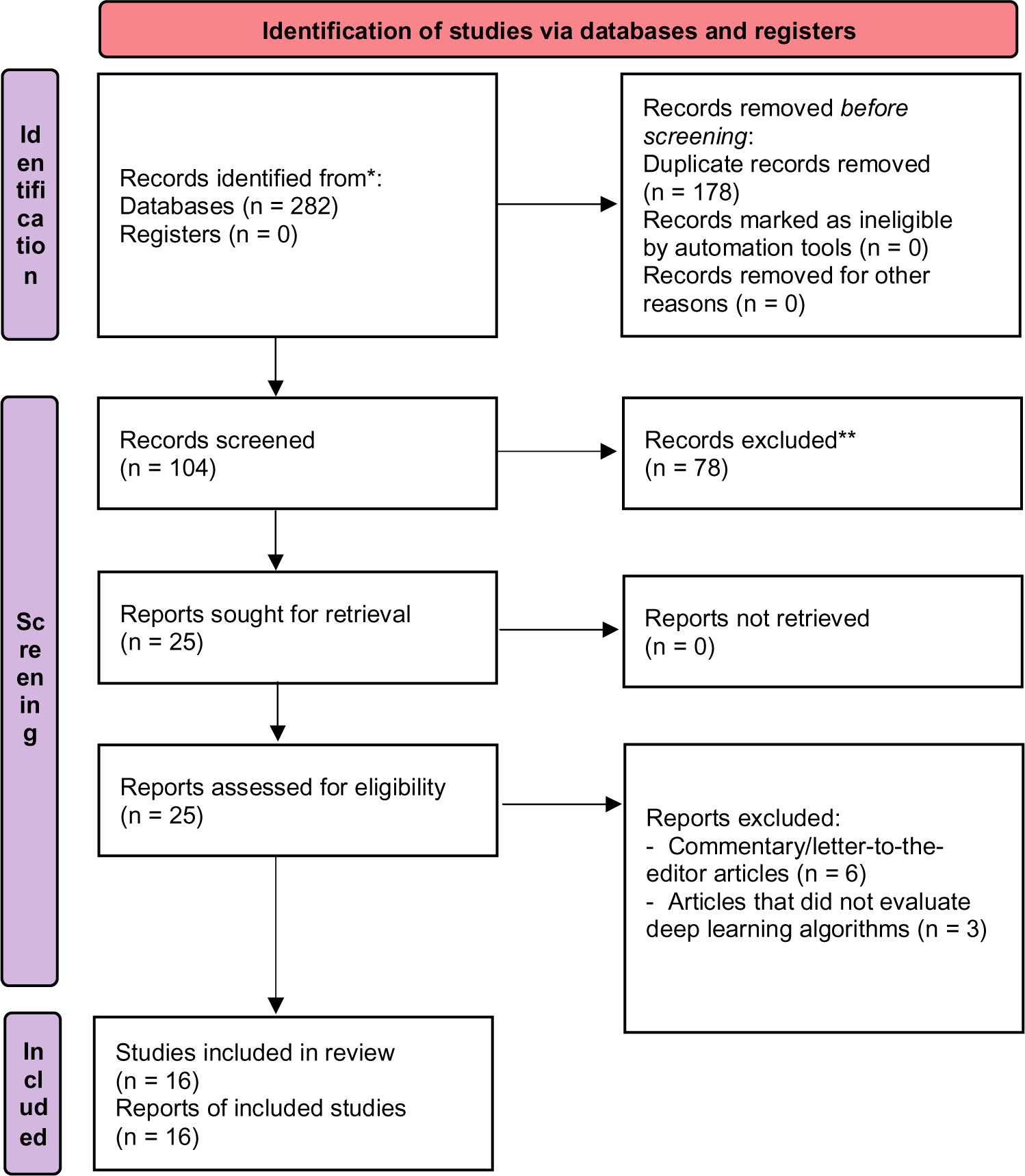
Flow Diagram of the inclusion process. Flow diagram of the search and inclusion process based on the Preferred Reporting Items for Systematic Reviews and Meta-Analyses (PRISMA) guidelines.

### Screening and Synthesis

This review was reported according to the PRISMA (Preferred Reporting Items for Systematic Reviews and Meta-Analyses) guidelines. Two reviewers (VS, EK) independently implemented the search strategy. The reviewers screened the titles and reviewed the abstracts of the articles identified in the search. The reviewers then assessed the full text of the relevant studies. Results of the studies included were summarized in a table format and included the title, journal, year of publication, study design, size of dataset, and performance metrics.

### Performance Characteristics

Performance characteristics for DL algorithms are evaluated based on sensitivity (also called recall), specificity, area under receiver operating characteristic curve (AUC), precision (also positive predictive value), accuracy and F1 score. The reported performance metrics from each study were summarized in a Table.

The quality of the studies was evaluated based on the Quality Assessment of Diagnostic Accuracy Studies (QUADAS-2) criteria(9).

## Fundamental Concepts

### Contrast-Enhanced Mammography (CEM)

*CEM* is an advanced breast imaging modality that combines the use of iodine-based contrast agent with conventional digital mammography (4). With CEM, dual energy for mammographic acquisition after IV administration of iodine-based contrast agent, is used. Two sets of images are obtained: images with low-energy exposures, and images at high-energy exposures, just above the kVp of iodine. The two exposures are subtracted, and two images are generated for each projection: low-energy images that are analogues to standard 2D mammography, and subtracted contrast-enhanced images displaying areas of contrast enhancement (**Figure 2**) (10).

**Figure 2.**
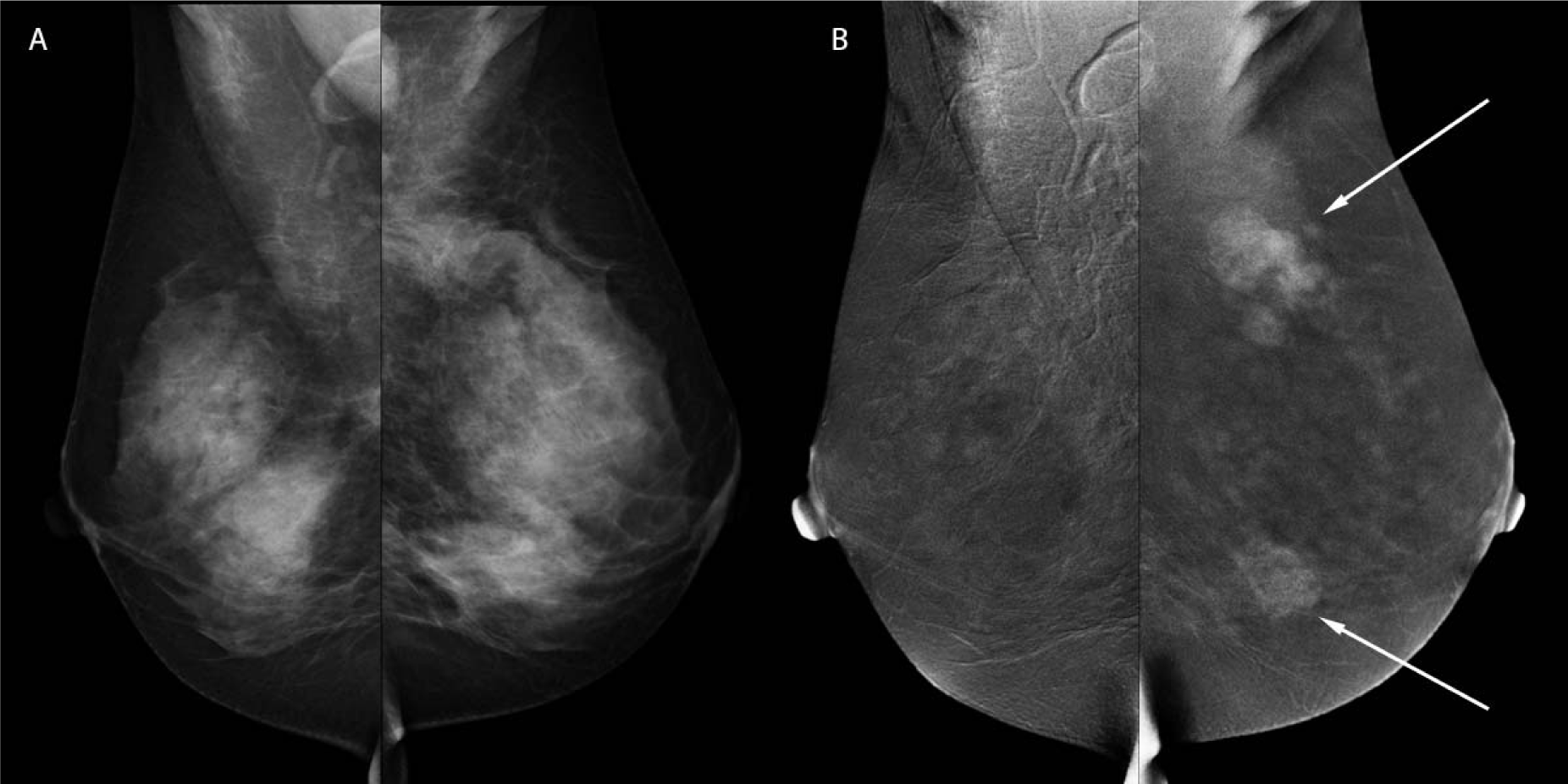
Contrast-enhanced mammography. An example of a contrast-enhanced mammography examination in a 45-year old woman. Mediolateral oblique low-energy (A) and subtracted contrast-enhanced (B) images of both breasts show multiple enhancing masses in her left breast. Ultrasound-guided biopsies were performed, confirming multifocal invasive ductal carcinoma (IDC).

*Background parenchymal enhancement (BPE)* refers to the normal enhancement of breast glandular elements and is graded on a BI-RADS scale from 0 to 3, indicating minimal, mild, moderate, or marked enhancement (**Figure 3**) (11). BPE at CEM correlates with age, menopausal status, breast density, and with BPE seen at MRI (12, 13). It is also associated with increased risk for breast cancer (13). More importantly, it may affect the diagnostic accuracy of CEM, as small abnormalities may blend with BPE when it is increased (11).

**Figure 3.**
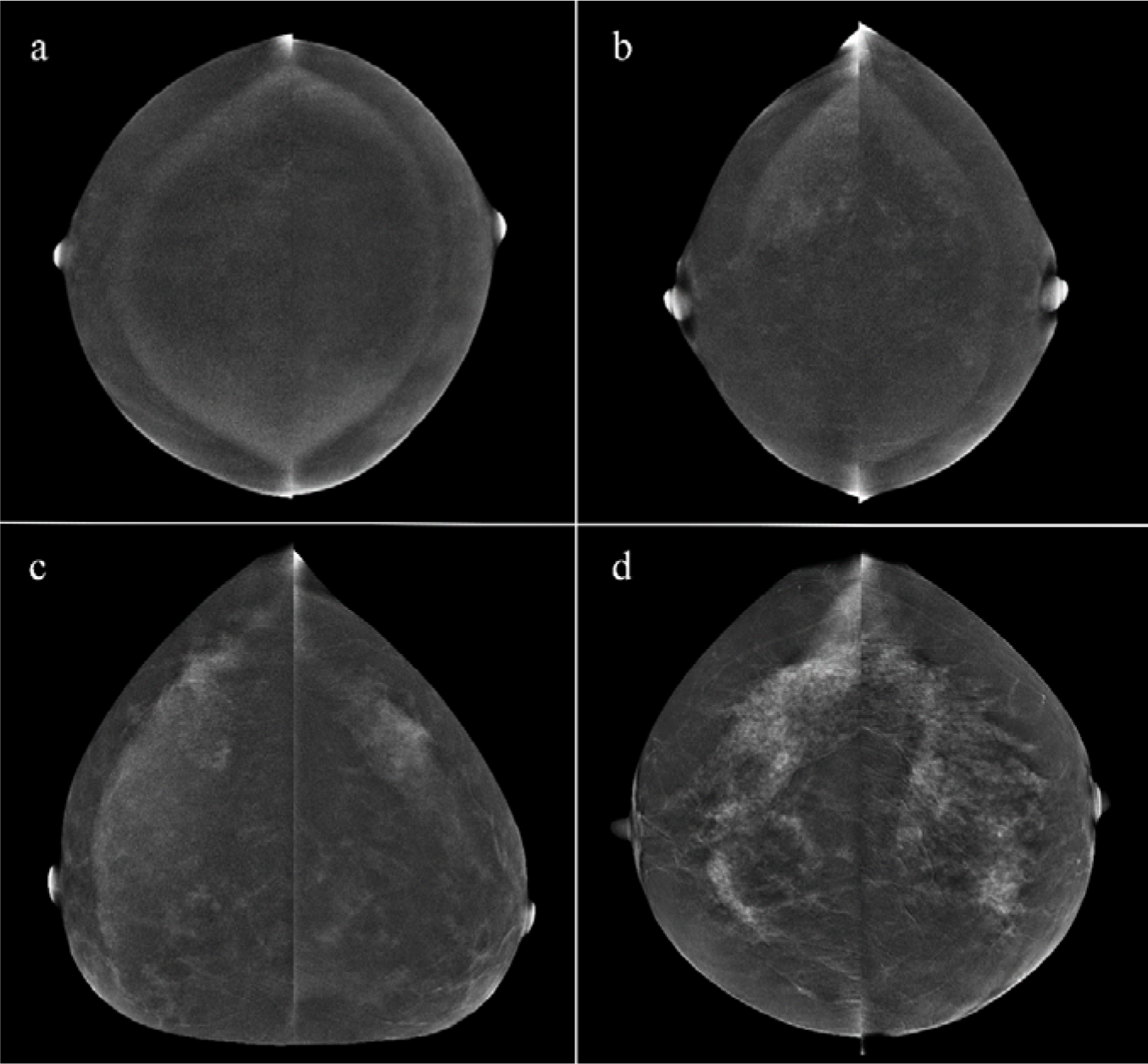
Background parenchymal enhancement (BPE) at contrast-enhanced mammography. Examples illustrating BPE grading at contrast-enhanced mammography: (a) Minimal BPE, < 25%; (b) Mild BPE, 25-50%; (c) Moderate BPE, 50-75%; (d) Marked BPE, > 75%. Reprinted with permission from Sorin et al. *Background Parenchymal Enhancement at Contrast-Enhanced Spectral Mammography (CESM) as a Breast Cancer Risk Factor*. Academic Radiology, 2020.

### Models and Techniques

*Deep Learning* represents a subset of machine learning (**Figure 4**) that employs neural networks with multiple layers to analyze complex relationships in data. It excels in identifying patterns and making decisions with minimal human intervention. In the domain of medical imaging, DL algorithms process extensive imaging data, learning to analyze pathological findings with precision that often matches or exceeds that of expert radiologists. For instance, a DL model can identify malignant tumors in CEM images. This capability may not only enhance diagnostic accuracy, but also streamline the workflow, thus impacting patient outcomes through appropriate intervention (7, 14).

**Figure 4.**
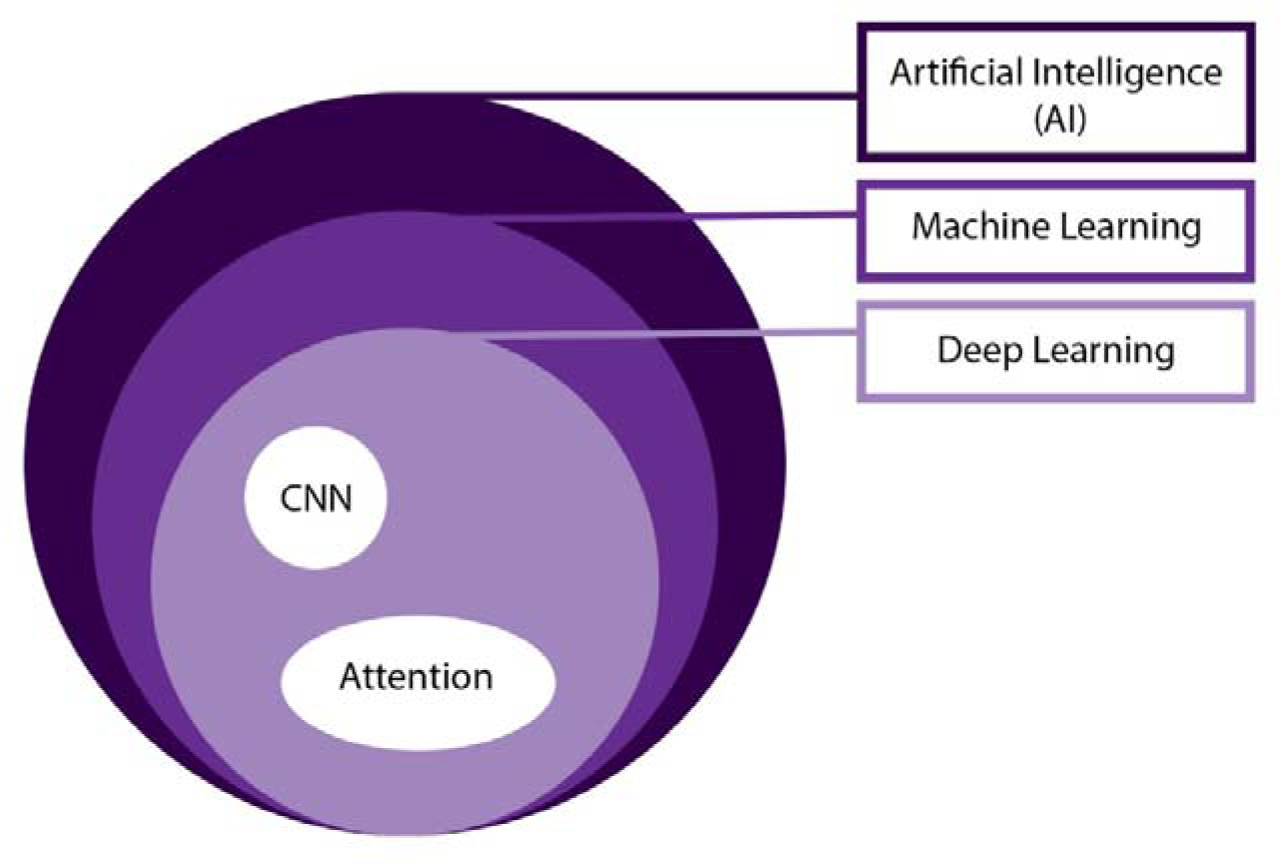
Venn diagram of artificial intelligence frameworks. This diagram illustrates that artificial intelligence (AI) is the broadest category, encompassing machine learning and deep learning as its subsets.

*Multi-Layer Perceptron (MLP)* is a neural network model that consists of at least three layers of artificial neurons: an input layer, hidden layers and an output layer (15). MLP utilizes backpropagation with supervised learning for training. With supervised learning the algorithm is provided with examples of inputs with correct outputs. During backpropagation the network adjusts its internal parameters whenever it makes an error. That way, it improves its accuracy over time (15).

*Convolutional Neural Networks (CNNs)* are a cornerstone of deep learning in image analysis (7). These models are designed to automatically and adaptively learn spatial hierarchies of features from images. CNNs consist of multiple layers of convolutions with learnable filters, and fully connected layers, making them adept at processing 2D and 3D images. In breast imaging, CNNs can analyze CEM images to identify patterns associated with breast cancer. The architecture of CNNs enables them to focus on local features within an image, such as the shape, size, and margins of lesions, facilitating detailed analyses (7).

*Attention* is a relatively novel technology that was widely evaluated in DL natural language processing (NLP) (16). In fact, the novel large language models (LLMs) such as GPT are based exactly on this mechanism (17). With image analysis algorithms, attention can improve their performance by allowing the models to focus on the most relevant features within an image (18). This is similar to the way that humans focus on specific parts of an image to extract information.

*Multimodal Networks* integrate and process data from multiple sources or types, for example text and images (**Figure 5**) (19). This approach is highly relevant with medical data analysis, as patient care relies on diverse data sources such as clinical history, lab tests, imaging etc.

**Figure 5.**
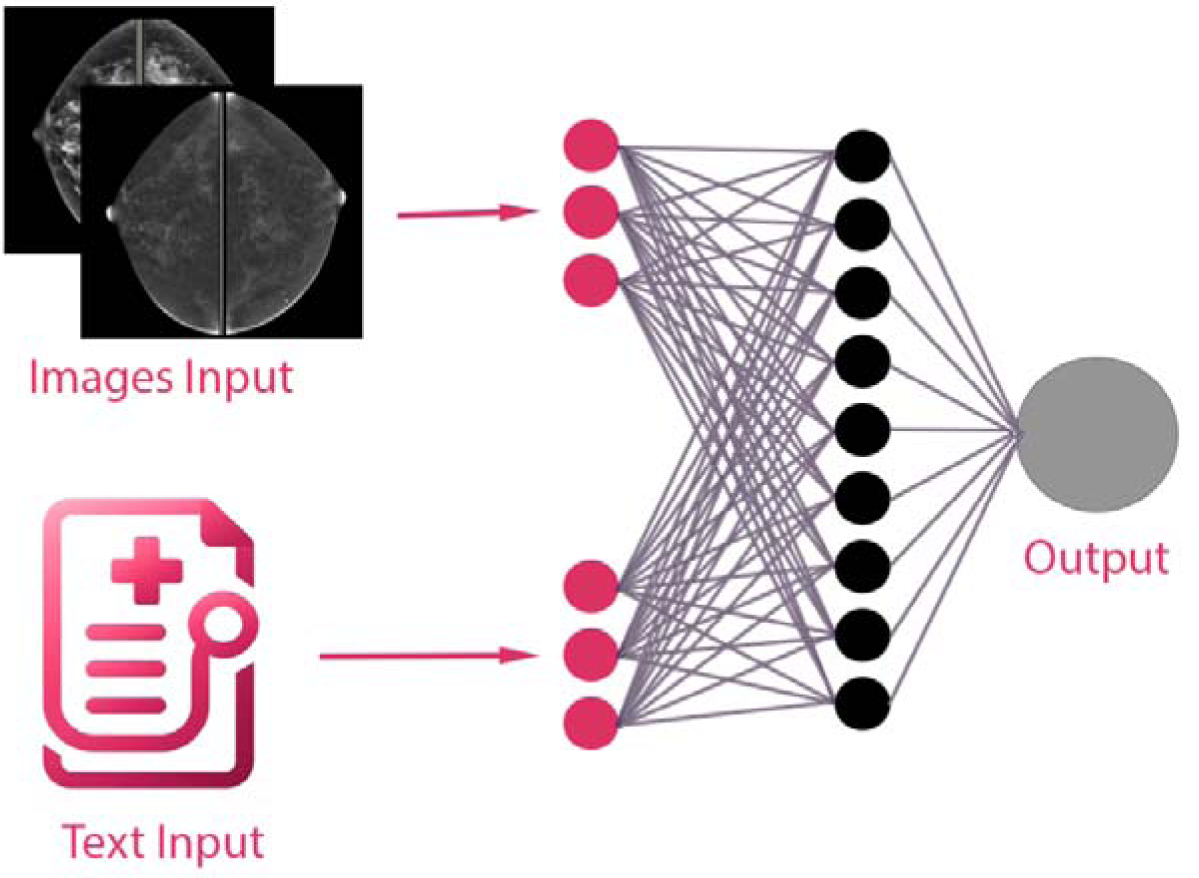
An illustration of a multimodal deep learning model. This diagram demonstrates the integration of a clinical note (textual data) and contrast-enhanced mammography images (visual data) within a neural network architecture. These are processed through distinct pathways before being merged in a unified framework, facilitating a comprehensive analysis in the context of patient diagnosis.

*Transfer Learning* is a technique used widely in DL models training. With this approach, a model developed for a specific task is reused as the starting point for a model applied for a different task. It transfers knowledge from the previously learned context to the new application, potentially enabling to achieve better performance with less data (**Figure 6**) (20).

**Figure 6.**
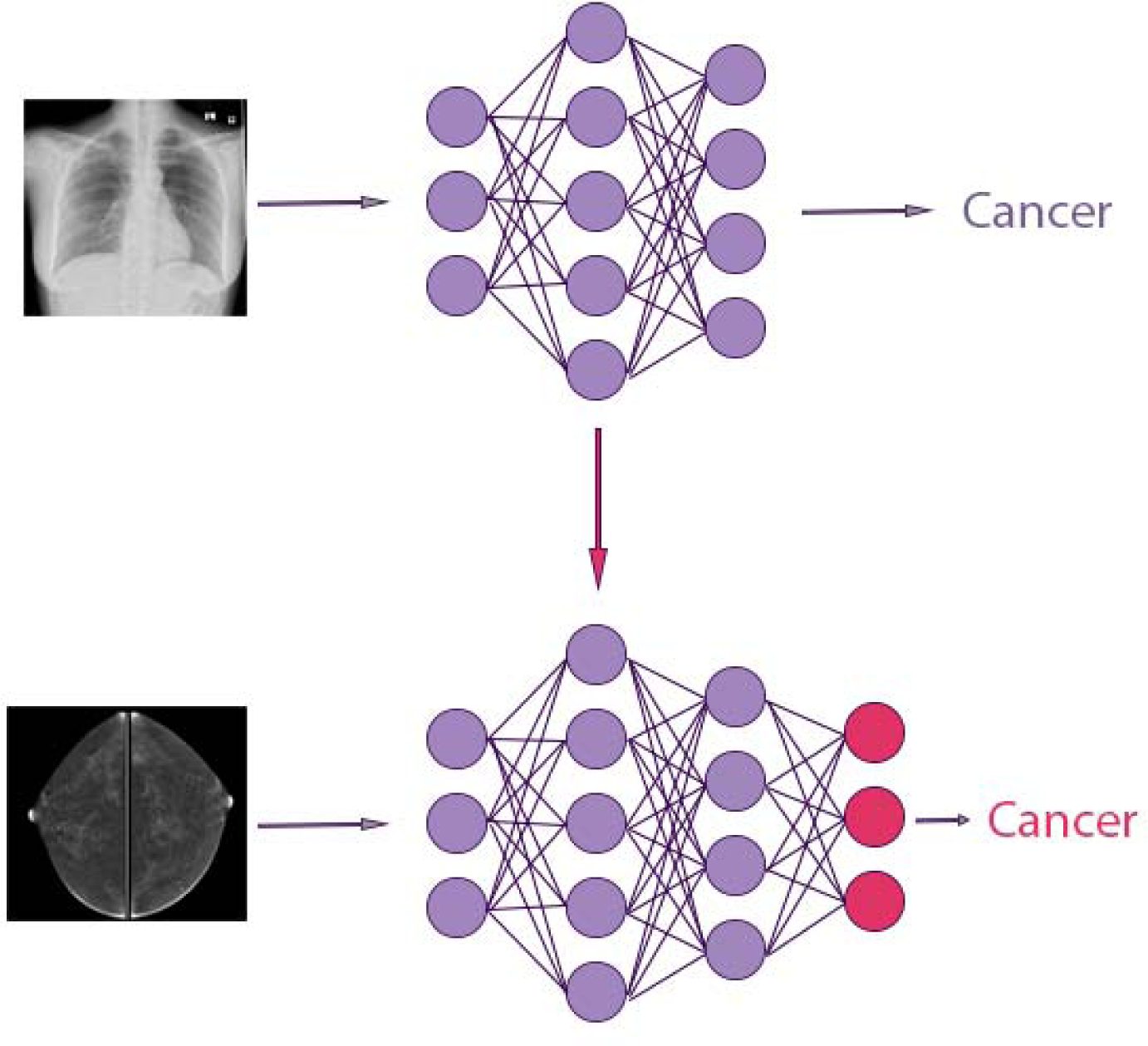
Illustration of transfer learning. This diagram displays the application of transfer learning between two neural network models. Initially, a chest X-ray image is processed by the first model, which is used for lung cancer classification. An arrow then signifies the transfer of learned features (in purple) to a second model, which processes a contrast-enhanced mammography (CEM) image. This second model is adapted to the characteristics of the CEM data, utilizing modified or additional components (in pink) for cancer classification at CEM.

*Generative Adversarial Network (GAN)* is a type of DL model that is aimed at generating new images (21). The model is composed of a generator creating fake images from input data, and a discriminator attempting to distinguish between fake and real images. The process enables both networks to improve over time through a competitive process where the generator aims to fool the discriminator (**Figure 7**) (21).

**Figure 7.**
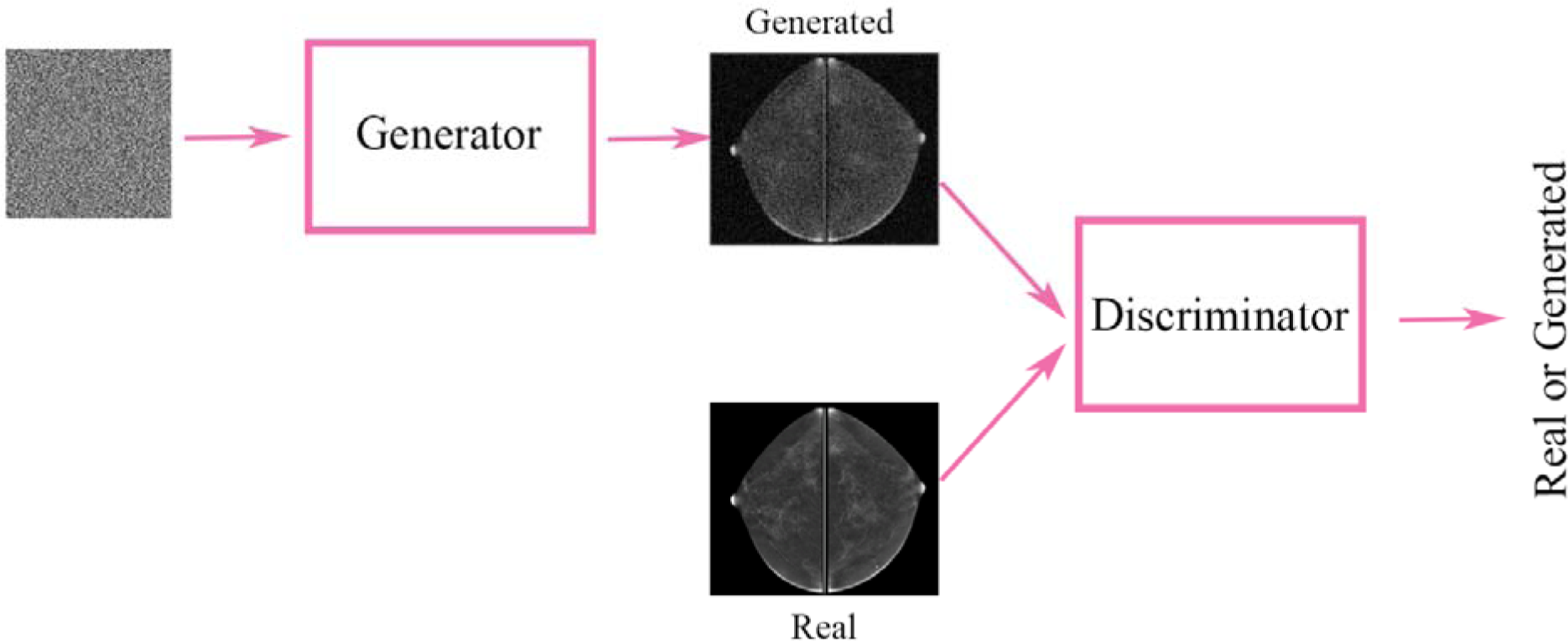
Schematic model of a generative adversarial network (GAN). The diagram illustrates how the generator and discriminator components are trained simultaneously. Starting with random noise as input, the generator produces synthetic images. These images, along with actual images, are supplied to the discriminator. The discriminator assigns a probability indicating whether the image is real. Based on feedback from the discriminator the generator then adjusts its parameters to enhance the realism of the synthetic images.

### Computer Vision Tasks

In medical imaging three foundational computer vision tasks play pivotal roles: classification, detection, and segmentation **(Figure 8**). Each of these tasks leverages DL algorithms to enhance diagnostic accuracy (7, 14).

**Figure 8.**
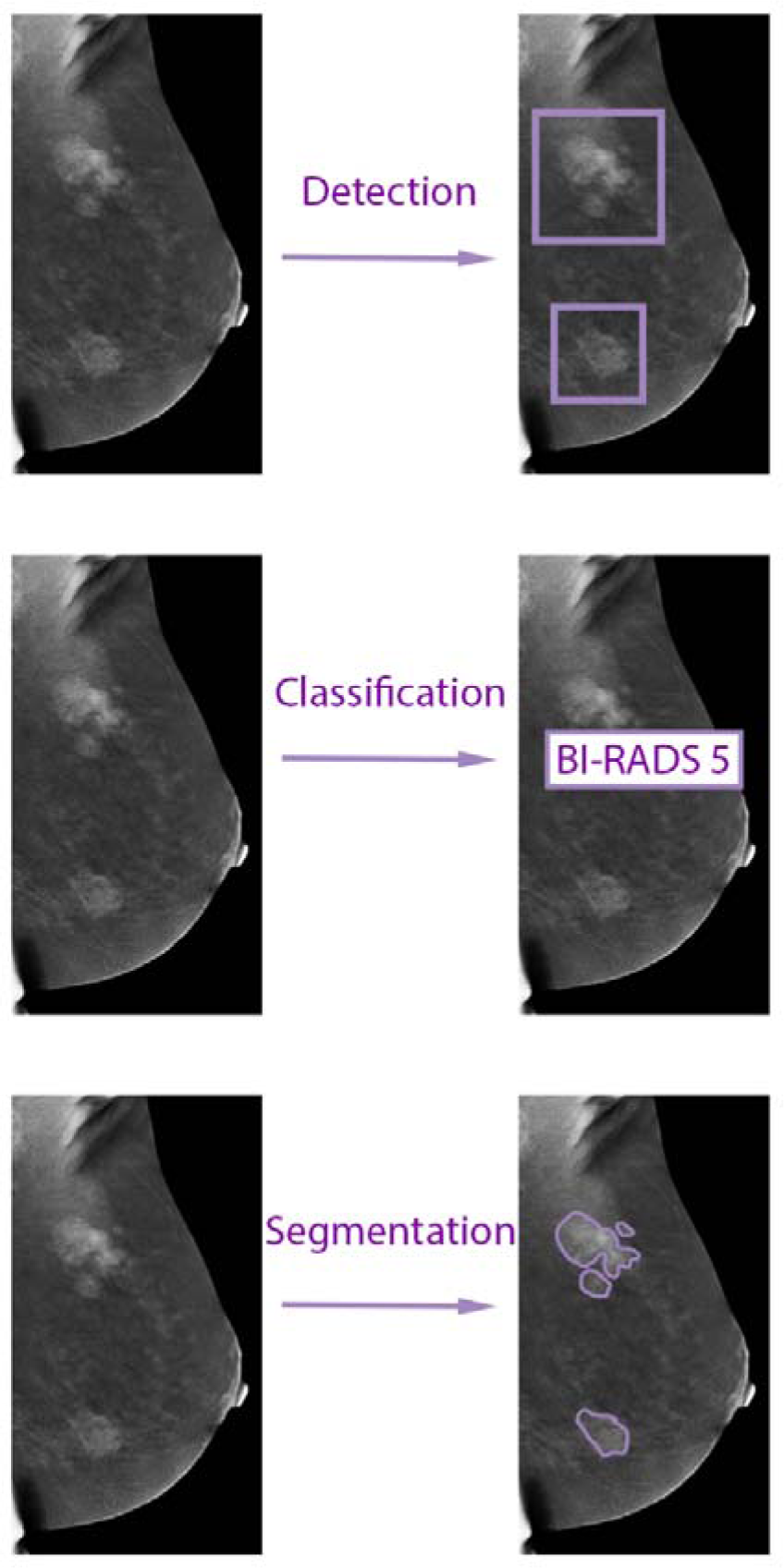
Image analysis tasks in medical imaging. This figure illustrates three tasks applied to contrast-enhanced mammography (CEM) images: lesion detection, classification, and segmentation.

*Detection* refers to the identification and localization of lesions within the images. For example, using DL algorithms to mark any abnormalities at CEM images with a circle or a box region of interest (ROI). This may aid radiologists in focusing their examination on areas of interest, potentially revealing lesions that may have been overlooked.

*Segmentation* is the process of delineating the exact boundaries of a lesion, separating it from the surrounding tissue. With manual segmentation, a radiologist traces the contours of a lesion in a CEM image, a time-consuming task with variability between observers.

Automated segmentation on the other hand, uses DL to precisely outline the lesion, facilitating a more consistent and efficient analysis. This is also valuable in assessing the size and shape of a tumor, which are critical factors in diagnosis and treatment planning. U-Net is a commonly used CNN segmentation network in the medical domain (22).

*Classification* involves the categorization of lesions into predefined classes, such as benign or malignant. For instance, in CEM, a DL model might analyze an image to classify a detected lesion as BI-RADS 2 (benign, requires routine screening), BI-RADS 3 (probably benign, requires short-term follow-up) or as BI-RADS 4 (potentially malignant, and requires tissue sampling).

By integrating DL algorithms into these computer vision tasks, researchers and clinicians can significantly improve the diagnostic process in breast imaging.

## Results

Sixteen studies have been included in this review, published between 2018 and 2024. While all the studies but one used CNN models, two of the CNN studies augmented the algorithm with attention layers.

All studies evaluated DL algorithms for classification of lesions at CEM, while six studies also assessed lesion detection or segmentation (**Figure 8**). In three studies segmentation was performed manually, two studies evaluated both manual and automatic segmentation, and nine studies automatically segmented the lesions. The objective assessment of the risk of bias based on QUADAS-2 is reported in **Table 1**. The results of the studies are summarized in **Table 2**.

**Table 1:** Quality Assessment of Diagnostic Accuracy Studies-2 (QUADS-2)

| *First Author* | **Patient Selection** | **Index Test** | **Reference Standard** | **Flow and Timing** | **Patient Selection** | **Index Test** | **Reference Standard** |
| --- | --- | --- | --- | --- | --- | --- | --- |
| *Helal et al.* | ✅ | ✅ | ✅ | ✅ | ✅ | ✅ | ✅ |
| *Chen et al.* | ✅ | ✅ | ✅ | ✅ | ✅ | ✅ | ✅ |
| *Jailin et al.* | ⚠ | ✅ | ✅ | ✅ | ? | ✅ | ✅ |
| *Qian et al.* | ✅ | ✅ | ✅ | ✅ | ✅ | ✅ | ✅ |
| *Beuque et al.* | ✅ | ✅ | ✅ | ✅ | ✅ | ✅ | ✅ |
| *Zheng et al.* | ✅ | ✅ | ✅ | ✅ | ⚠ | ✅ | ✅ |
| *Li et al.* | ⚠ | ✅ | ✅ | ✅ | ? | ✅ | ✅ |
| *Mao et al.* | ✅ | ✅ | ✅ | ✅ | ⚠ | ✅ | ✅ |
| *Wu et al.* | ✅ | ✅ | ✅ | ✅ | ⚠ | ✅ | ✅ |
| *Dominique et al.* | ✅ | ✅ | ✅ | ✅ | ⚠ | ✅ | ✅ |
| *Khaled et al.* | ⚠ | ⚠ | ✅ | ✅ | ? | ? | ✅ |
| *Song et al.* | ⚠ | ✅ | ✅ | ✅ | ? | ✅ | ✅ |
| *Song et al.* | ⚠ | ⚠ | ✅ | ✅ | ? | ? | ✅ |
| *Perek et al.* | ⚠ | ⚠ | ✅ | ✅ | ? | ? | ✅ |
| *Gao et al.* | ⚠ | ⚠ | ✅ | ✅ | ? | ? | ✅ |
| *Danala et al.* | ⚠ | ⚠ | ✅ | ✅ | ? | ? | ✅ |

**Table 2.**
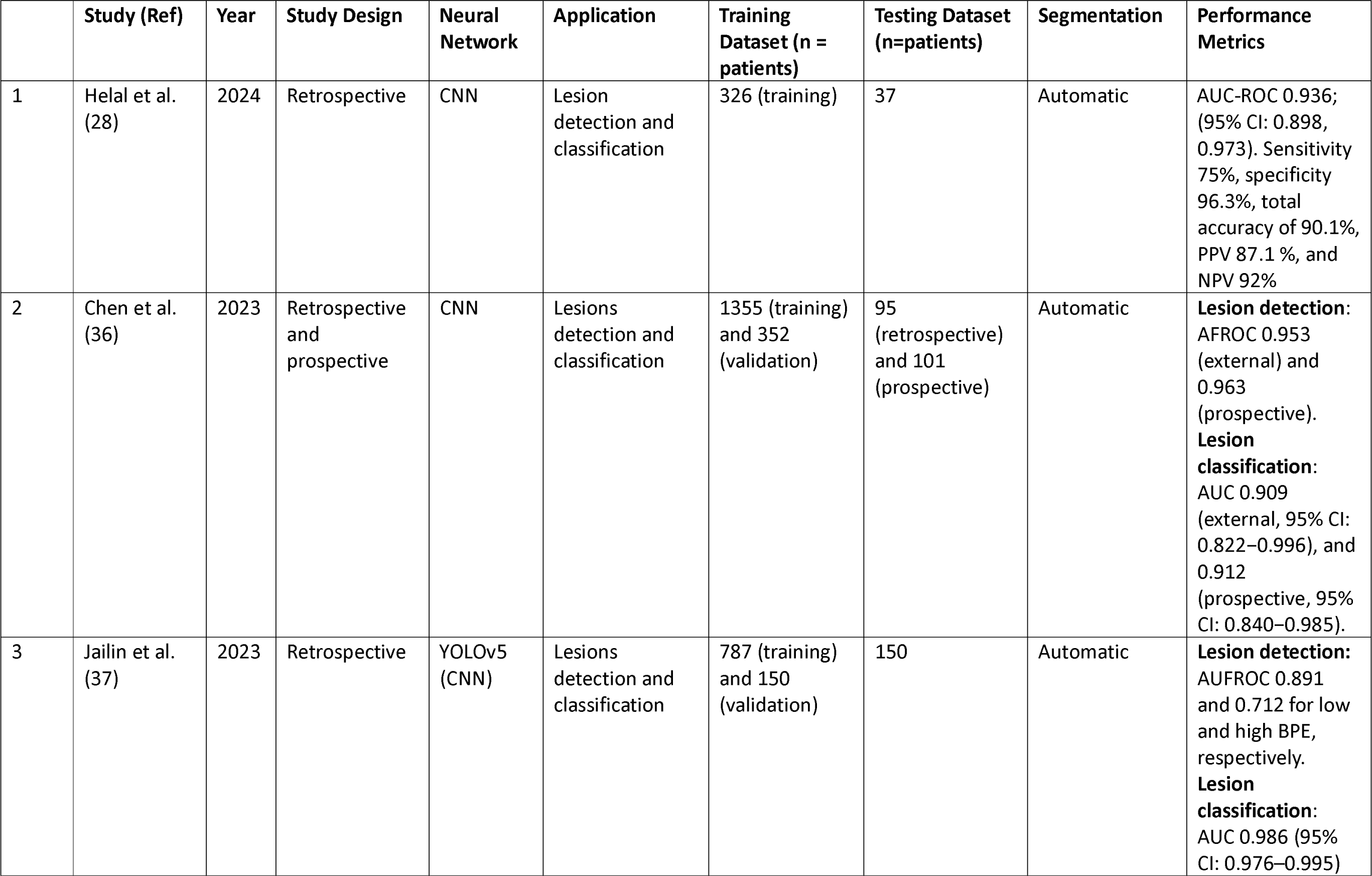
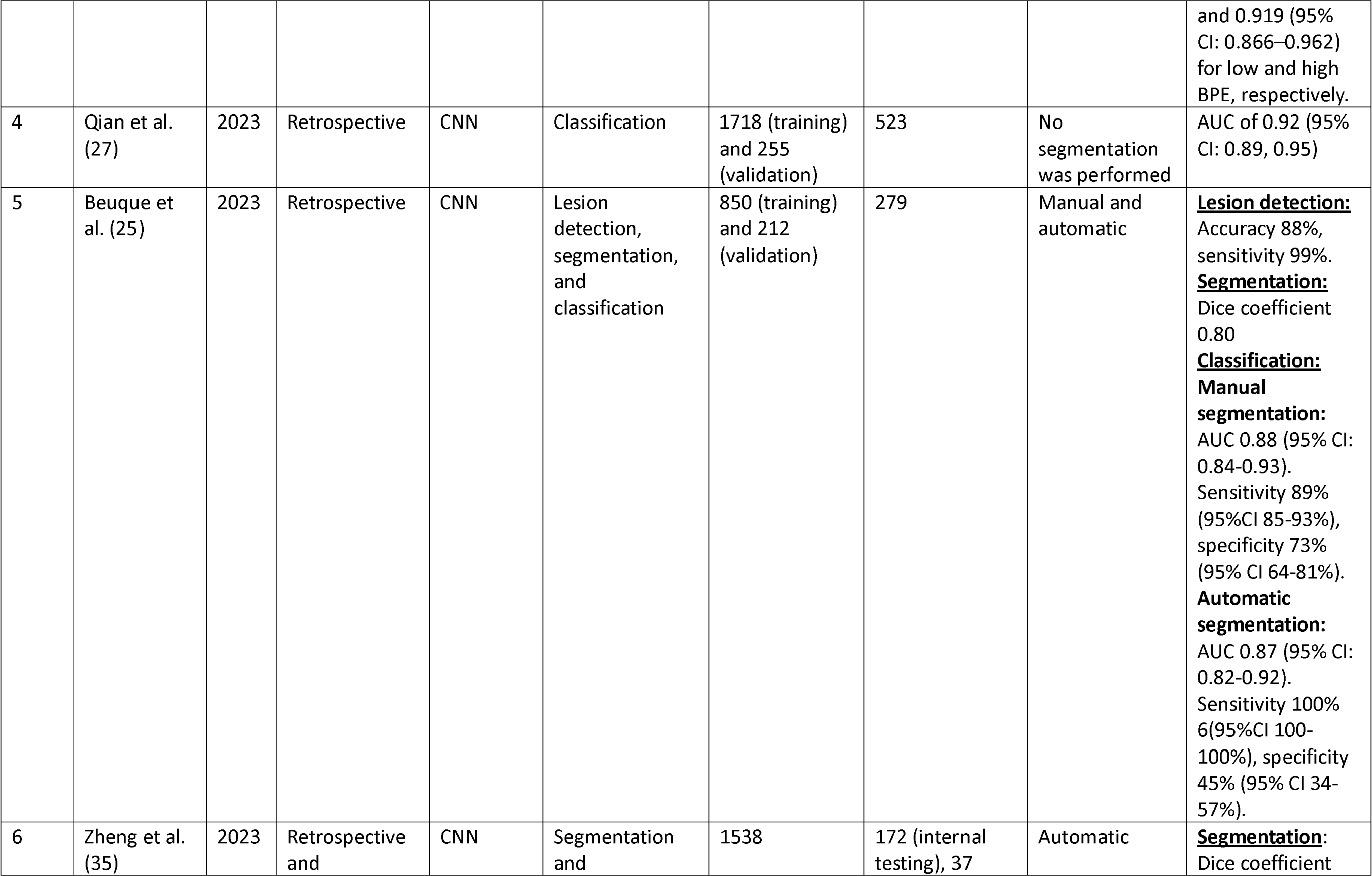
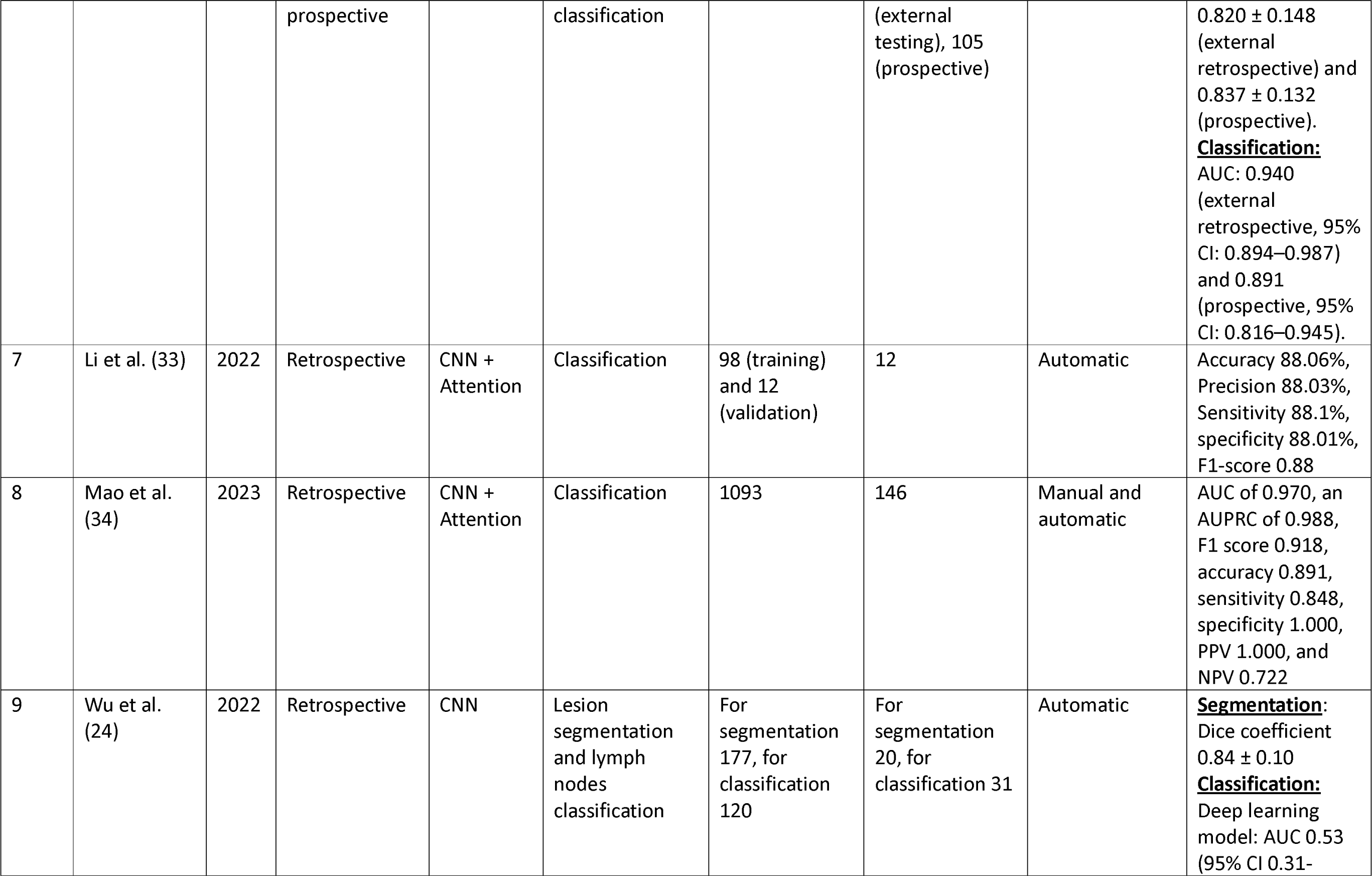
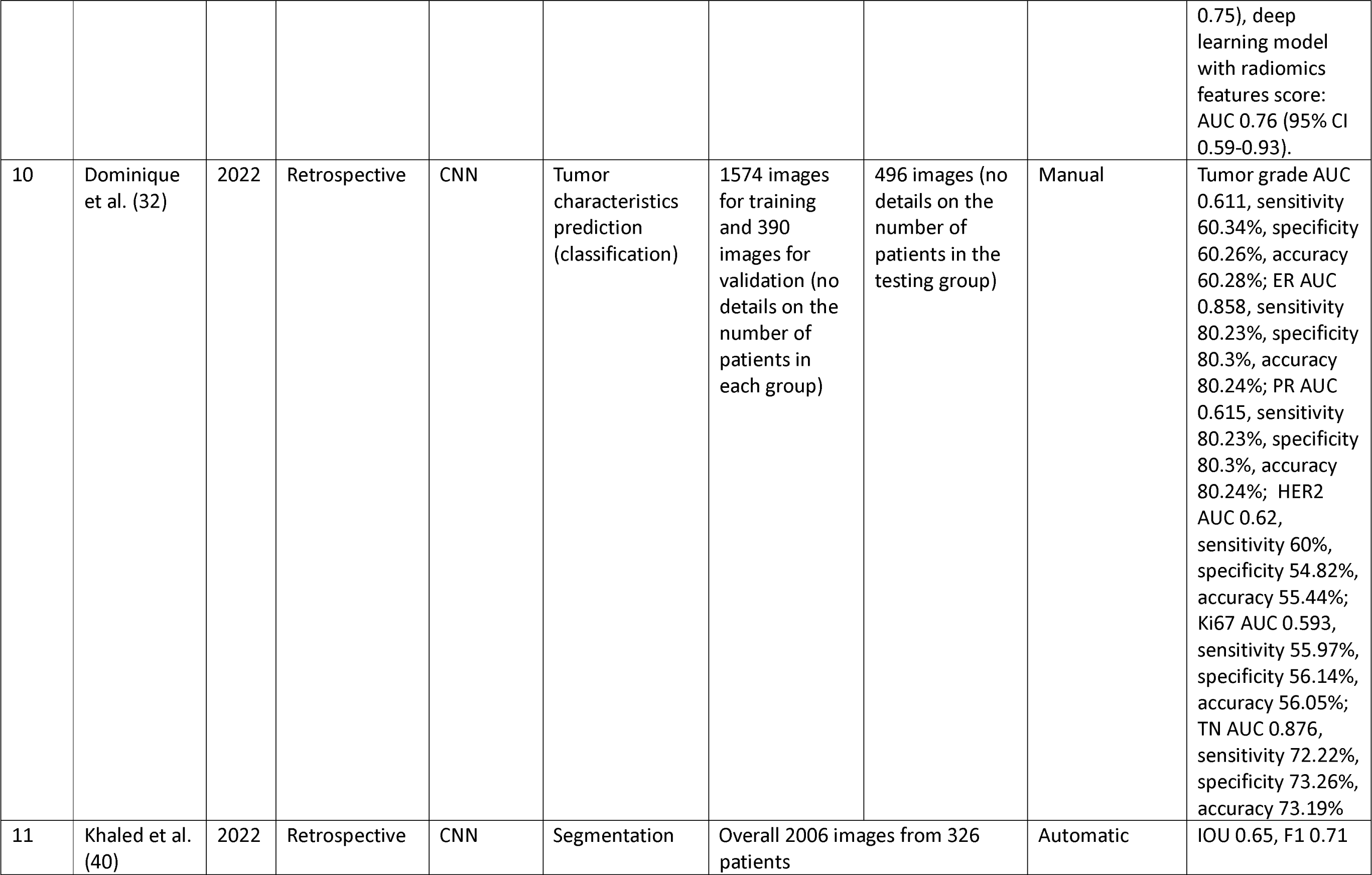
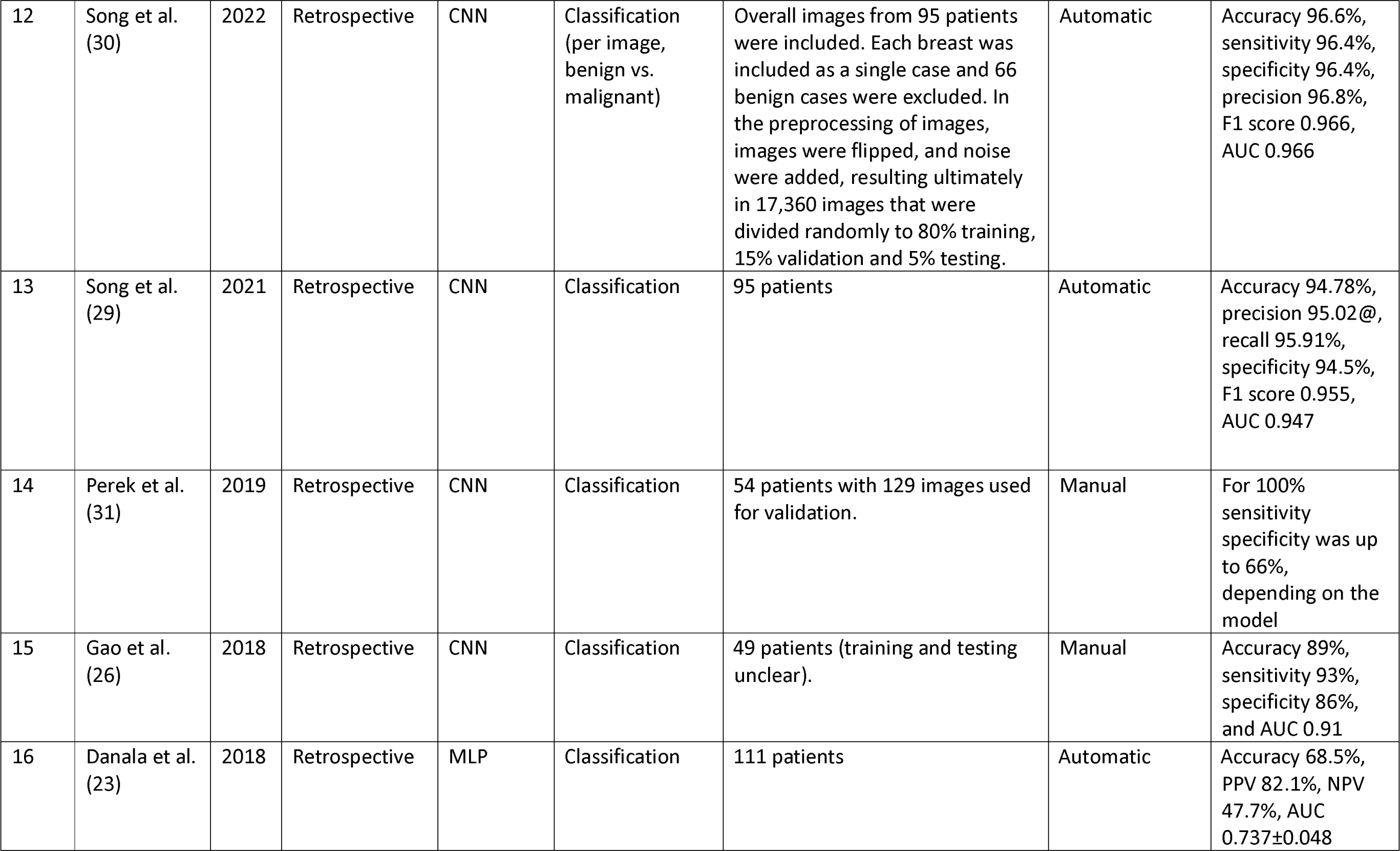
Studies Evaluating Deep Learning Applications for Contrast Enhanced Mammography.

### Segmentation

Danala et al. (23) were the first to publish on DL analysis of CEM images. They developed MLP for lesion segmentation at CEM, and classification between malignant and benign lesions. They were the first to use automatic segmentation of lesions based on lesion enhancement in the subtracted contrast images. Then, based on these segmentations they analyzed both the subtracted and the low-energy images for lesion classification (23).

Wu et al. (24) combined DL models with a radiomics model. Radiomics involves the extraction of a large number of quantitative features from images. The authors first used a U-Net, which is a supervised DL network, for image segmentation. Training was performed on manually segmented images. Then, using another DL network (ResNet-18) they extracted features from the segmented tumor regions of interest in both the low-energy and the subtracted contrast images. They’ve also incorporated radiomics features to predict non-sentinel lymph node metastases. The radiomics model without DL overperformed the DL, and the combined model (24). Beuque et al. (25) developed a full-workflow model for automatic lesion identification, segmentation, and classification. A radiomics model was also trained to classify both manual and automatically segmented lesions. The DL model had the highest sensitivity for cancer classification, while the combined DL with radiomics model had higher specificity with the highest AUC (25).

### Detection and classification

Gao et al. (26) developed a CNN model that classifies breast lesions based on features from both low-energy and recombined contrast-enhanced images. They have also applied a CNN model that learned the mapping between the low-energy and recombined images, to generate “virtual” contrast-enhanced images from standard digital mammography images. Using features from these “virtual” recombined images, the performance of the model in lesion classification improved, compared to incorporating only features from standard digital mammography (26). Qian et al. (27) have developed a multi-feature fusion neural network to combine information from both the low-energy and the contrast-enhanced images, for breast cancer detection.

In a recently published study, Helal et al. (28) also developed a Multiview DL model analyzing both low-energy and subtracted contrast images for breast lesions classification at CEM studies and compared its performance to radiologists. The DL model had lower sensitivity for breast cancer detection compared to radiologists, while specificity for the model was higher (28). In 2021 Song et al. (29) evaluated a CNN that analyzed both the low-energy images and the subtracted contrast-enhanced images simultaneously to classify lesions. Later, in 2022, they’ve developed a model that applies GAN (**Figure 3**) used for fusion of the subtracted contrast-enhanced images and the low-energy images, and the classification model for lesion classification (30).

Perek et al. (31) applied a multimodal network for lesion classification in the subtracted CEM images, combining textual features with image analysis. They compared fine-tuning a pretrained neural network (AlexNet) and fully training a CNN. Their multimodal network resulted in a theoretical reduction in benign biopsies by up to 66% (31).

Dominique et al. (32) fine-tuned an existing CNN model that was used for chest X-rays analysis, on CEM dataset. They applied the model to predict histopathological characteristics of breast cancer from CEM images. For example, receptor status, grade and proliferative index. The model achieved the best results for estrogen receptor status prediction from the subtracted contrast-enhanced images (32).

Two studies that assessed the combination of an attention mechanism with CNN (33, 34). First, Li et al. (33) incorporated a CNN algorithm with attention for CEM images classification. Their network extracts information from all four images acquired for each breast (low energy and subtracted contrast images). In their study they report that this method significantly improved the accuracy of cancer classification and reduced false-positive cases compared to other DL models previously reported. Later, Mao et al. (34) also developed an attention-based DL model to discriminate benign from malignant breast lesions on CEM images. They’ve evaluated and compared between three CNNs with attention, and the same models without attention. The attention-based models outperformed those without attention mechanism. They have also compared the best model performance to that of a radiomics model, as well as to radiologists. They show that the radiologists’ performance improved with the algorithm’s support (34).

Prospective data analysis validates the performance of DL models in real-world scenarios. It ensures these models are robust and applicable to future, unseen cases, beyond retrospective data. We found only two studies that evaluated their model on prospective data. Zheng et al. (35) developed a fully automated pipeline system based on CEM images for segmentation and classification of breast lesions. They assessed the algorithm as a support tool for breast radiologists, demonstrating improved sensitivity and specificity in a prospective testing cohort (35). Chen et al. (36) also developed a multi-process model for detection and classification of breast lesions at CEM. They have assessed their model in a retrospective external testing set as well as a prospective dataset. The model outperformed senior breast radiologists in lesion classification metrics (36).

Jailin et al. (37) developed a DL algorithm for detection and classification of breast lesions using data from both the low-energy and subtracted contrast images. They then evaluated their algorithm’s performance at different levels of BPE (**Supplemental Figure 3**), showing increased false positive rates in cases with increased BPE (37).

## Discussion

CEM is a novel imaging technique that has higher sensitivity compared to standard 2D mammography for detecting breast cancer, albeit with a decrease in specificity. We demonstrated that DL algorithms can be used for CEM lesion detection, segmentation and classification. Notably, some of the models have showed superior accuracy compared to radiologists’ evaluations. Furthermore, integrating models as supportive tools for CEM interpretation has refined specificity while keeping high sensitivity, and decreased the false-positive rate.

While the integration of DL for CEM analysis is still at a relatively early stage, there are studies that developed full-process applications that are able to detect, segment and classify breast lesions (25, 28, 35–37). Despite the promising results, currently, there is an absence of FDA-approved AI algorithms available for CEM (38).

Integrating DL algorithms into clinical workflows has the potential to enhance breast cancer screening and diagnosis. The algorithms can enable more accurate study interpretations in less time. Notably, the increased specificity reported for some algorithms highlights their utility in using these models as support tools for CEM studies interpretation. This could diminish the frequency of recalls and biopsies, while maintaining high sensitivity. Large, prospective clinical trials are necessary to assess the clinical value and real-world effectiveness of utilizing DL algorithms as a support tool for reading CEM studies. The ongoing prospective MASAI trial in Sweden is an example, assessing the application of DL in the interpretation of standard screening mammography (39).

The integration of AI into clinical workflows is anticipated to affect the role of breast radiologists. It may enable radiologists to dedicate more attention to complex cases, while spending less time on straightforward examinations. Radiologists may have higher availability for involvement in multidisciplinary teams. Initial automatic analysis of cases may allow triage and prioritization of examinations based on urgency. The transformation ultimately may streamline patient management, enhance the timeline of medical interventions, and promote a patient-focused approach in breast cancer care. Ultimately, implementation of these algorithms in everyday clinical use could improve patient outcomes and overall efficiency of healthcare delivery.

This review has several limitations. First, heterogeneity of studies and variability in measures between studies prevented a meta-analysis. Second, DL in radiology and CEM are both rapidly expanding topics. Thus, there may be relevant studies published after our review was performed. Finally, this review did not fully account for the variation in clinical settings and patient populations across the studies analyzed. This variability can affect the generalizability of the findings, as the performance of the algorithms could differ based on the study indications and patient demographics. Such differences might influence the algorithms’ effectiveness and applicability in diverse contexts.

In conclusion, CEM is an emerging modality in breast imaging with a high diagnostic performance, outperforming standard 2D mammography and comparable to breast MRI. At present, the application of DL to CEM is at its beginning but showing promising performance. When used as a support tool, DL can potentially improve radiologists’ accuracy and efficiency in the interpretation of CEM examinations, improving specificity while keeping high sensitivity for cancer detection. Further research is needed to validate DL models across different practices and CEM vendors, and prospectively assess their impact on real-world clinical practice.

## Data Availability

All data produced in the present work are contained in the manuscript

